# Benchmarking Multimodal Large Language Models for Forensic Science and Medicine: A Comprehensive Dataset and Evaluation Framework

**DOI:** 10.1101/2025.07.06.25330972

**Authors:** Ashmaan Sohail, Om M. Patel, Jihwan Choi, Jack C. S. Venditti, Addison J. Wu

## Abstract

**Background:** Multimodal large language models (MLLMs) have demonstrated substantial progress in medical and legal domains in recent years; however, their capabilities from the lens of forensic science—a field that is at the intersection of complex medical reasoning and legal interpretation, with conclusions critiqued by judicial scrutiny—remains largely unexplored. Forensic medicine uniquely depends on the accurate integration of often ambiguous text and visual information, yet systematic evaluations of MLLMs in this setting are lacking.

**Methods:** We conducted a comprehensive benchmarking study of eleven state-of-the-art MLLMs, including proprietary (GPT-4o, Claude 4 Sonnet, Gemini 2.5 Flash) and open-source (Llama 4, Qwen 2.5-VL) models. Models were evaluated on 847 examination-style forensic questions drawn from various academic literature, case studies, and clinical assessments, covering nine forensic subdomains. Both text-only and image-based questions were included. Model performance was assessed using direct and chain-of-thought prompting, with automated scoring verified through manual revision.

**Results:** Performance improved consistently with newer model generations. Chain-of-thought prompting improved accuracy on text-based and choice-based tasks for most models, though this trend did not hold for image-based and open-ended questions. Visual reasoning and complex inference tasks revealed persistent limitations, with models underperforming in image interpretation and nuanced forensic scenarios. Model performance remained stable across forensic subdomains, suggesting topic type alone did not drive variability.

**Conclusions:** MLLMs show emerging potential for forensic education and structured assessments, particularly for reinforcing factual knowledge. However, their limitations in visual reasoning, open-ended interpretation, and forensic judgment preclude independent application in live forensic practice. Future efforts should prioritize the development of multimodal forensic datasets, domain-targeted fine-tuning, and task-aware prompting to improve reliability and generalizability. These findings provide the first systematic baseline for MLLM performance in forensic science and inform pathways for their cautious integration into medico-legal workflows.

## Introduction

Large language models (LLMs) are artificial intelligence systems that are capable of generating human-like textual responses to prompts through processing vast amounts of text data. Recent studies have shown that LLMs can nearly match the precision of diagnosis and treatment selection to that of specialized physicians. These models, including GPT-4o, Claude 4 Sonnet, Meta Llama 4 Maverick, and other proprietary and open-source variants, have become more capable at a variety of tasks over time ^1–3^. Notably, this advancement of LLMs has now been showcased across a range of societally-embedded knowledge-intensive fields, including medicine, education, and law, often evaluated through standardized examinations ^4–9^.

Despite this progress, little research has explored how LLMs can be integrated into forensic science, particularly forensic medicine, which is a unique field encompassing the intersection of both human medicine and law. Existing studies focus heavily on the analysis of legal documents, clinical decision support, and other medical examinations, each task being investigated in isolation from one another ^4,7,9^. Forensic science presents unique challenges for artificial intelligence applications ^10,11^. As a cornerstone of public safety and justice systems, knowledge of forensic science is invaluable in informing investigations that can lead to societally impactful decisions. Unlike other medical fields, where diagnostic protocols and evidence-based treatments are navigated through comprehensive patient histories, forensic science often relies on the interpretation of ambiguous data within complex legal and judicial frameworks, where conclusions are critiqued by judicial scrutiny ^12–16^.

A differentiating aspect of forensic medicine is its especially heavy reliance on both visual and textual information. Tasks such as injury pattern recognition, postmortem changes assessment (lividity, decomposition stages, etc.), or trace evidence evaluation (fibres, ballistic markings, etc.) depend heavily on imaging ^17–20^. Vision-language models (VLMs) extend on LLMs to process both text and image input, thereby being essential to the advancement of AI applications in this field ^21–24^. Notably, the systematic evaluations of VLMs in this field are nearly non-existent.

Moreover, existing evaluations of foundation models in medicine primarily relied on structured, single-source, text-based examinations, such as medical licensing tests or board certifications ^25– 33^. Forensic assessments, however, extend beyond factual recall or clinical decision-making and are highly variable in format, often incorporating case-based scenarios, applied knowledge, and visual interpretation ^12,15,16^. There is a clear need to understand how VLMs perform when exposed to complex, multimodal forensic investigations to determine their role in education, assessment, and practice.

In this study, we evaluated the performance of eleven state-of-the-art open-source and closed-source multimodal LLMs (MLLMs) on a collection of 847 examination-style questions. Rather than relying on a single standardized test, we aggregated questions from publicly available academic resources and case studies in an attempt to reflect the real-world diversity of forensic assessments. We compared their accuracy, analyzed the ability of MLLMs in medical reasoning, legal and clinical guidelines, and other multimodal scenarios. To our knowledge, this is the first benchmarking study evaluating MLLM performance across a comprehensive, multimodal forensic science dataset. Specifically, our objective is to address the following questions:

a. How do different MLLMs perform when answering different types of questions about forensic science?
b. What are the potential applications and current limitations of using MLLMs to support forensic science education, assessment, and real-world integration?

## Methodology

### Section I: Dataset

To benchmark the capabilities of various VLMs in forensics medicine, we developed a diverse question bank of 847 total questions, covering a variety of topics.

We relied upon publicly available educational sources to construct this dataset. These included leading undergraduate and graduate-level forensics science textbooks, such as Forensic Pathology Review by Wayne, Schandl, and Presnell; and Forensic Science: From the Crime Scene to the Crime Lab by Saferstein ^16,34^. We also obtained copies of nationally-certified observed structured clinical examinations in forensic science by the University of Jordan Faculty of Medicine ^35^.

Our dataset spans nine representative topics in forensic science: death investigation and autopsy; toxicology and substance usage; trace and scene evidence; injury analysis; asphyxia and special death mechanisms; firearms, toolmarks, and ballistics; clinical forensics; anthropology and skeletal analysis; and miscellaneous/other. As shown in Figure 1, the most represented topics in the dataset were death (n = 204), toxicology (n = 141), trace (n = 133), and injury (n = 124). Topics such as asphyxia and special death mechanisms (n = 70); firearms, toolmarks, and ballistics (n = 60); clinical forensics (n = 49); and anthropology and skeletal analysis (n = 38) had moderate representation.

**Figure 1.**
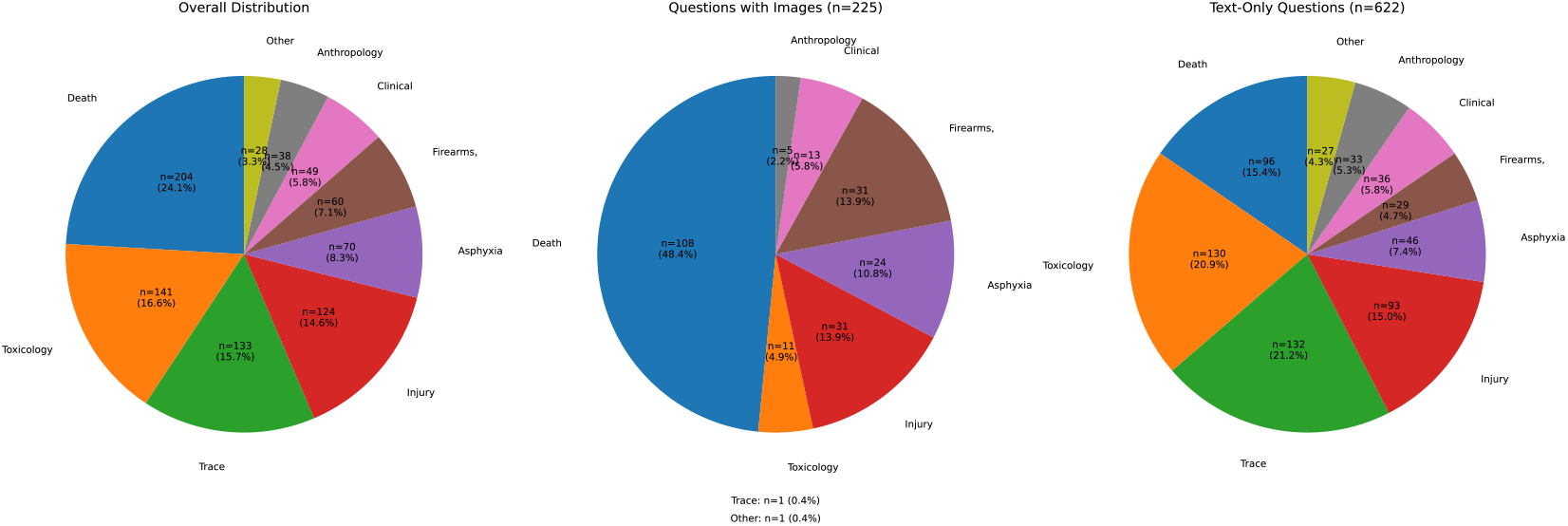
Distribution of topics of the curated dataset questions, segmented into all questions, image-based questions, and text-only questions.

Our dataset includes both text-only and image-based questions. Of the 847 total questions, 225 included an image (26.6%), while the remaining 622 (73.4%) were text-only. Image-based questions were most concentrated in the death investigation and autopsy category, which often require visual assessments of wounds, lividity, decomposition stages, and similar features. Text-based questions were most concentrated in the trace and toxicology section, which frequently involve interpreting chemical reports, identifying substances, and reasoning through forensic procedures based solely on written descriptions.

Several question formats were also included. Out of the 847 total questions, 781 questions (92.2%) were multiple choice questions with 4 or 5 option choices, or 2 – true and false, presented to the LLMs to choose from. The remaining 66 questions (7.8%) were non choice-based questions that often mimicked real-life forensic scenarios, requiring the LLMs to work through case narratives, interpret findings, and articulate conclusions in a manner reflective of professional forensic reporting or medico-legal consultation.

### Section II: Models and Evaluation

We evaluate a variety of frontier open-source and proprietary models and their respective predecessors. The proprietary vision-language models we evaluated in our study were GPT-4o, Claude 4 Sonnet, Claude 3.5 Sonnet, Gemini 2.5 Flash, Gemini 2.0 Flash and Gemini 1.5 Flash. The open-source vision-language models we evaluated in our study were Llama 4 Maverick 17B-128E Instruct, Llama 4 Scout 17B-16E Instruct, Llama 3.2 90B, Llama 3.2 11B, and Qwen2.5-VL 72B Instruct, which were all hosted on Together AI.

Both direct and chain-of-thought prompting were used in the evaluation of model performance of the question bank. In direct prompting, we steer the model to immediately provide its final answer without any intermediate thinking or reasoning. In contrast, in chain-of-thought prompting, we steer the model to reason about its thought process before providing its answer^36^. For all models, we set their temperatures to be their respective default throughout conducting all of the experiments to reflect a representative distribution of responses and capabilities. For the proprietary models – those in the OpenAI, Claude, and Gemini families – this is equivalent to a temperature of 1.0. For the open-source models hosted on together AI – those in the Llama and Qwen families – this is equivalent to a temperature of 0.7.

We score each question on a scale from 0 to 1, where a score of 0 indicates that the question is completely incorrectly answered, and a score of 1 indicates that the question is completely correctly answered. For single-part questions, the answer can either only be a 0 or a 1 as there is only one part to the question. For multiple-part questions, we weigh each part in the entire question equally, with the final score being the proportion of parts answered correctly. We award no partial credit for any part (either incorrect or correct). Automatic numerical evaluation was conducted by employing LLM-as-a-judge using GPT-4o for all model responses. To validate the accuracy of GPT-4o as an evaluator, we manually evaluated 30 randomly sampled responses from each model. For each sampled response, we compared the GPT-4o-generated score against a human annotation. We found perfect agreement between GPT-4o and human judgments across all samples, confirming its reliability as an automated evaluator in our setting ^37^.

## Results

For direct prompting, the accuracy observed for each language model exhibited considerable variation, ranging as low as 45.11% ± 3.27% for Llama 3.2 11B Vision Instruct Turbo to as high as 74.32% ± 2.90% for Gemini 2.5 Flash. When models are steered with chain-of-thought prompting, the accuracy range is higher overall, from an increased minimum of 51.0% ± 3.32% for Qwen 2.5-VL 72B, to an increased maximum of 79.0% ± 2.72% for Gemini 2.5 Flash. In all models tested, every model improved in accuracy when tested with chain-of-thought prompting compared to direct prompting (p < 0.005), with the single exception being Qwen 2.5-VL 72B, which demonstrated an 11.7% reduction in total accuracy when using chain-of-thought prompting (p < 0.001) (Fig. 2).

**Figure 2.**
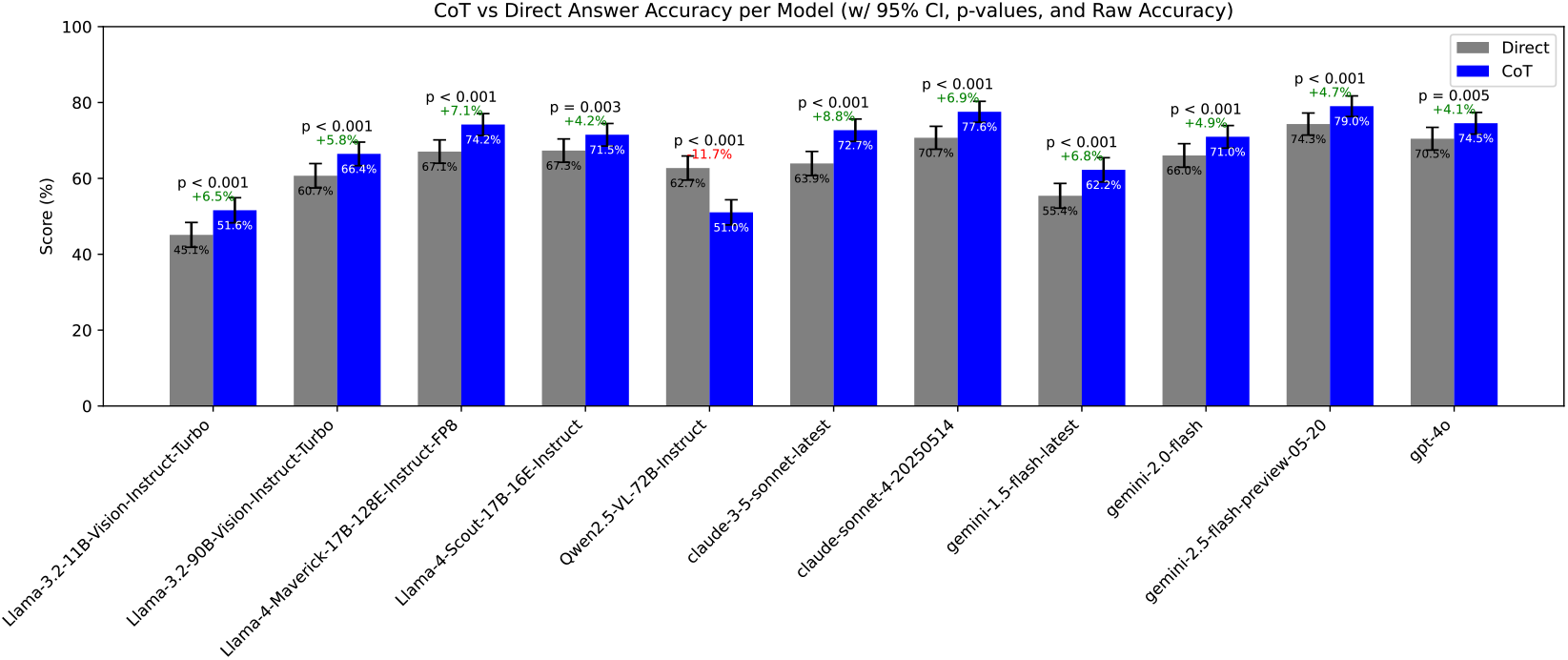
Aggregate accuracy for each model on the curated dataset, for both direct and CoT prompting.

Accuracy manifested a trend of improvement within model families when comparing older and newer versions. For the Gemini family models, we observed a 10.63% increase in accuracy (p < 0.001) for direct prompting and 8.72% (p < 0.001) for chain-of-thought prompting between Gemini 1.5 and 2. We also observed an increase in accuracy by 8.28% (p = 0.0001) for direct prompting and 8.07% (p < 0.001) for chain-of-thought prompting between Gemini 2 and 2.5 (Fig. 2).

Llama experienced a 15.58% increase (p < 0.001) between Llama 3.2 11B and Llama 3.2 90B, and a 6.37% increase (p = 0.0019) between Llama 3.2 90B and Llama 4 17B. Claude experienced a 6.78% increase (p = 0.0025) between Claude 3.5 and Claude 4 for direct prompting, and a 4.86% increase (p = 0.019) for chain-of-thought prompting. These results indicated that model age or generation was consistently linked with higher performance under both direct and chain-of-thought prompting. Furthermore, the data indicated that variation of the question topic did not significantly affect accuracy outcomes, as accuracy scores remained relatively uniform throughout. Accuracy remained broadly stable across forensic science topics (Fig. 3). To quantify topic-driven performance differences, we computed eta squared (η^2^) values from one-way ANOVAs per model, measuring the proportion of accuracy variance explained by topic. Across all models, η^2^ values ranged from 0.012 to 0.063, with most models well below the conventional threshold for a medium effect (η^2^ > 0.06). For example, GPT-4o yielded η^2^ = 0.046, Claude 3.5 Sonnet η^2^ = 0.063, and Qwen2.5-VL η^2^ = 0.012. These results indicate that the topic explained only a small fraction of accuracy variance, suggesting that model performance was largely consistent across the range of forensic subdomains included in the dataset and that the topic was likely not the primary factor for consistent performance changes.

**Figure 3.**
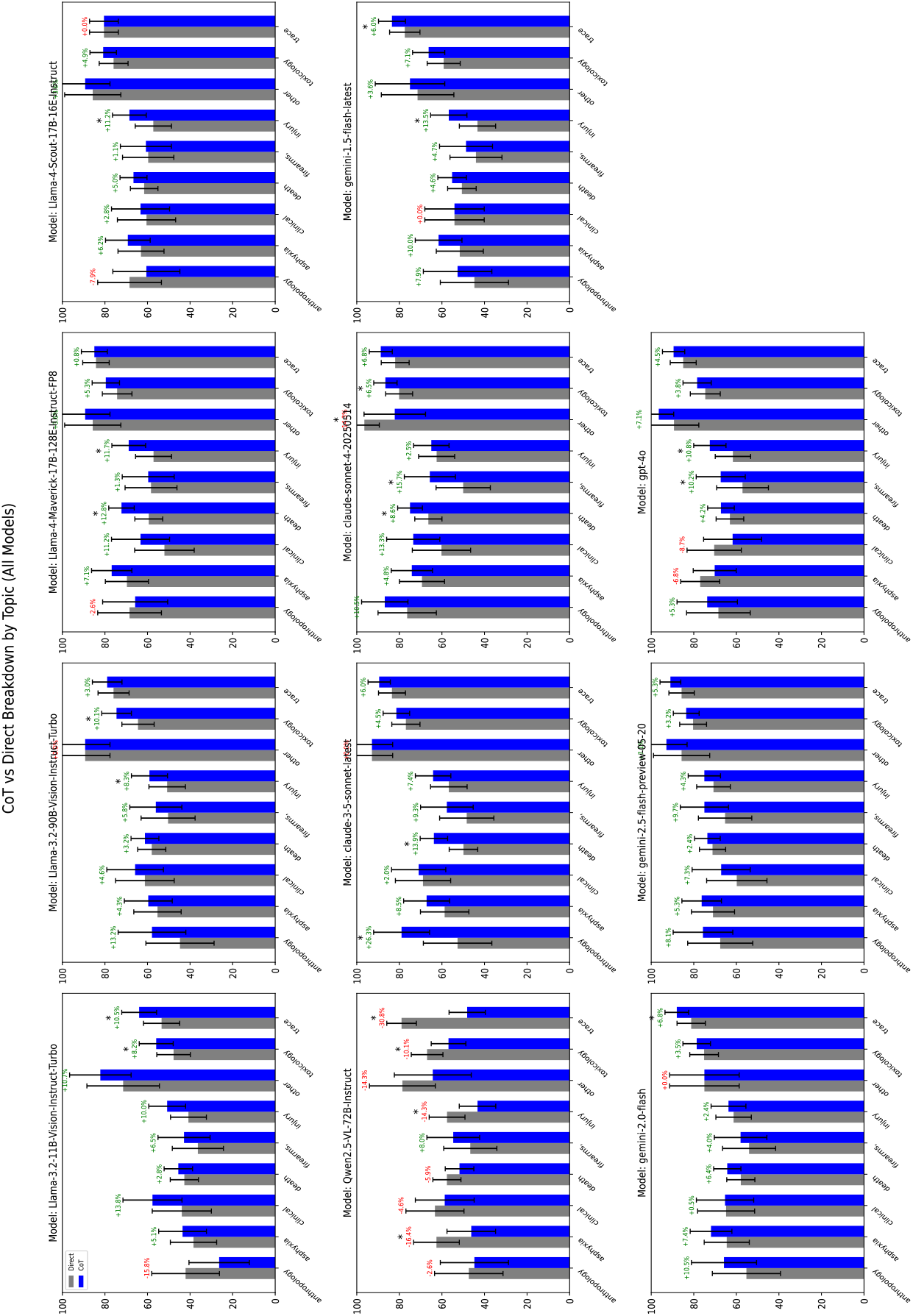
Performance for each model segmented into per-topic accuracy.

For text-based questions, chain-of-thought prompting improved accuracy for nearly all models (p

< 0.001; no statistically significant effect observed for GPT-4o p = 0.103) except for Qwen 2.5-VL 72B, which recorded a decrease of 15.1% (p < 0.001) when chain-of-thought prompting was used for text-based questions instead of direct prompting (Fig. 4).

**Figure 4.**
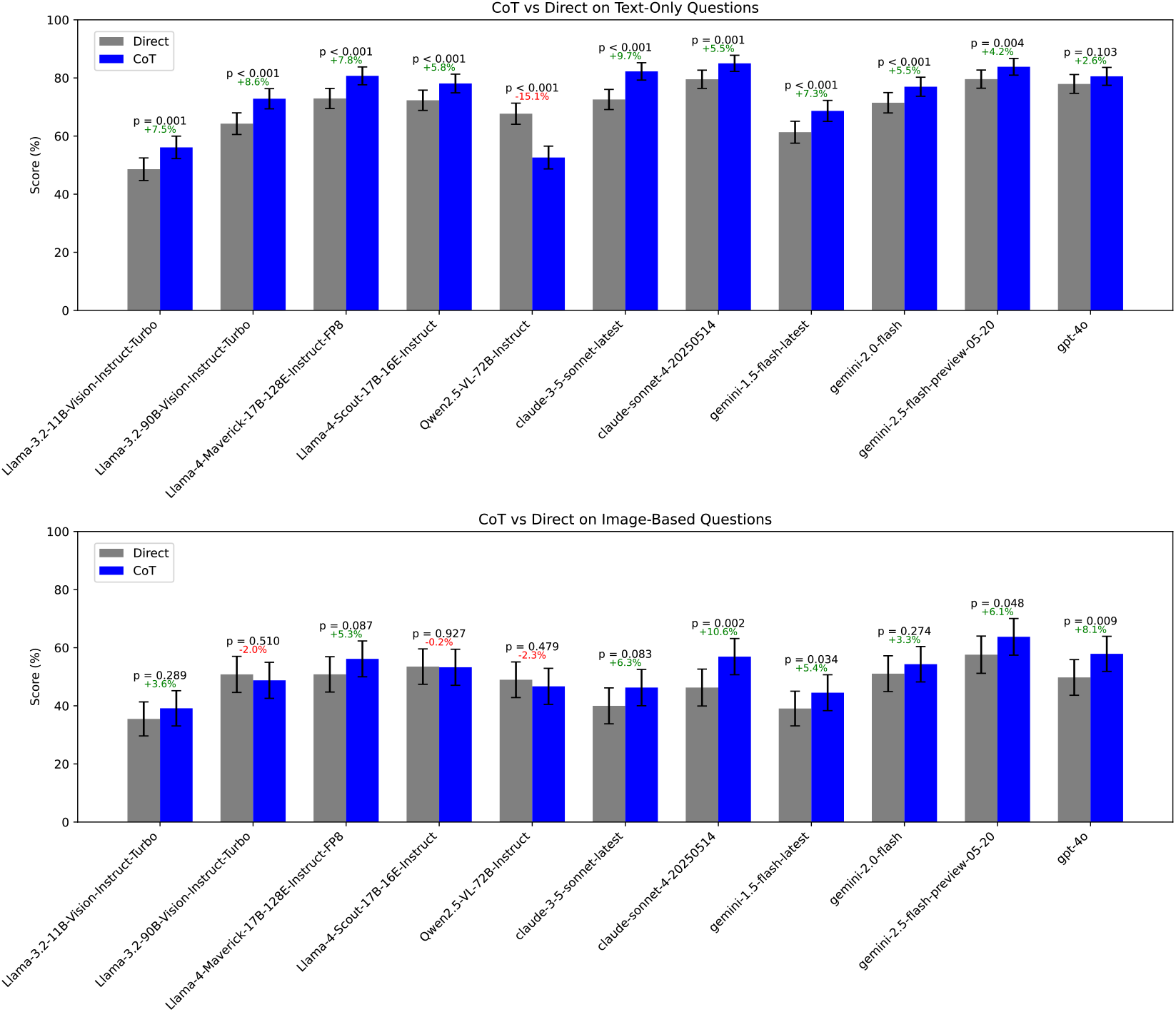
Performance of each model on text-only and image-based questions, for both direct and CoT prompting.

For image-based questions, we observed a different pattern in terms of the effectiveness of different prompting strategies. Compared to when models are evaluated on the text-based questions, we see fewer statistically significant gains in accuracy from chain-of-thought prompting compared to direct prompting. The only models that benefited from chain-of-thought prompting were Claude Sonnet 4 (p = 0.002), Gemini 1.5 Flash (p < 0.05), Gemini 2.5 Flash (p < 0.05), and GPT-4o (p < 0.01). Also, three models experienced decreased accuracy with chain-of-thought prompting on image-based questions, although none of these decreases were statistically significant. These models were Llama 3.2 90B, which experienced a 2% decrease, Llama 4 Scout 17B 16E, which experienced a .2% decrease and Qwen 2.5 72B which experienced a 2.3% decrease, each of which showed a lower accuracy when using chain-of-thought reasoning for image items than when using direct prompting alone (Fig. 4).

For choice-based questions, chain-of-thought prompting displayed consistent improvement for nearly all models tested, yielding an average increase of 6.1% relative to the results from direct prompting, with the exception of Qwen 2.5 72B, which demonstrated a 12.7% decrease. For non-choice-based questions, the results showed a reduced pattern. Most models recorded improvements when chain-of-thought prompting was applied; however, the Llama 3.2 90B showed a minor decrease in this area, with accuracy dropping by approximately 3.3% when chain-of-thought prompting was used instead of direct prompting for non-choice-based items. The average increase for non-choice-based questions across models was 4.99%, which was lower than the average improvement for choice-based questions (Fig. 5).

**Figure 5.**
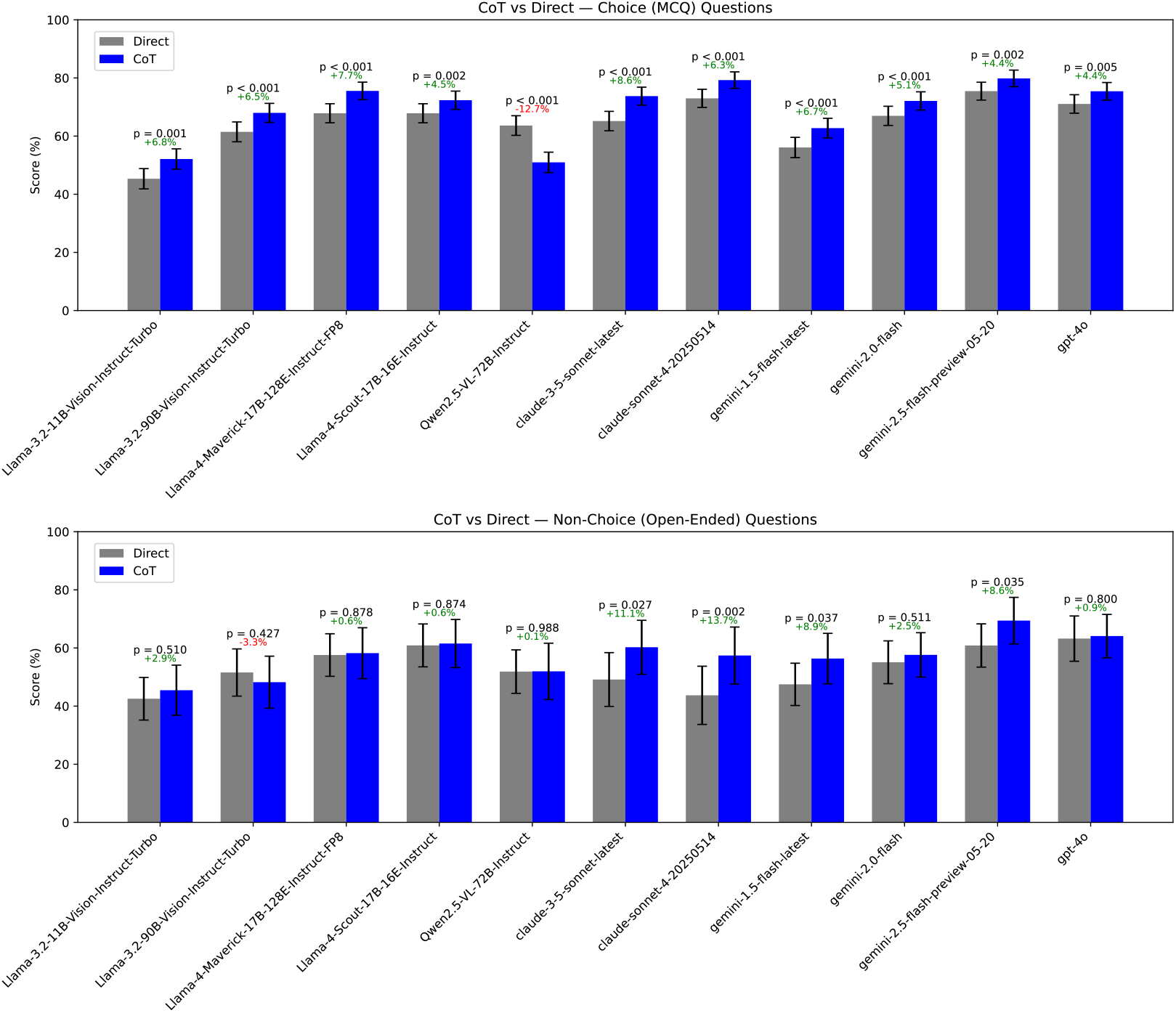
Performance of each model on choice-based and non-choice (open-response) questions, for both direct and CoT prompting.

We also noted that models consistently attained higher accuracy rates on text-based questions than image-based questions. The largest difference we observed was with the Claude 3.5 Sonnet model, with a text-based and image-based accuracy rate of 77.42% and 43.13% (p < .001), respectively. Furthermore, choice-based questions consistently attained higher accuracy rates than non choice-based questions for every model. The largest difference can be observed in the Claude Sonnet 4 model with a choice-based accuracy rate of 81.82% and a non choice-based accuracy rate of 50.56% (p < .001) (Fig. 5).

The results across all categories – direct versus chain-of-thought prompting, text-based versus image-based questions, and choice-based versus non-choice-based questions – consistently demonstrated measurable and interpretable differences. In general, accuracy improvements were greater for text-based and choice-based questions than for image-based and non-choice-based formats. The Qwen 2.5 72B model consistently performed with reduced accuracy in multiple categories when coupled with chain-of-thought prompting, whereas the other models presented consistent accuracy improvements on average. Within the same model families, newer models consistently performed better than older ones across the observed data. Finally, accuracy did not fluctuate significantly with variation of question topic, remaining relatively stable across the forensic science subjects included in the experiment.

## Discussion

In this study, to the best of our knowledge, we conducted the first systematic evaluation of state-of-the-art proprietary and open-source MLLMs on a novel forensic science dataset incorporating a variety of topics and modes of question delivery. Our results revealed clear performance trends and suggestions for improvement – accuracy generally improved with newer model generations and chain-of-thought prompting, but this benefit was primarily exclusive to text-based and choice-based tasks. Performance on image-based and open-ended questions remained inconsistent, highlighting persistent limitations in applying current MLLMs to real-world forensic scenarios.

Our study has several limitations. While the dataset we derived is diverse and drawn from credible academic and clinical literature, the distribution of topics was not balanced. Questions from several subfields such as forensic entomology, forensic odontology, and forensic toxicokinetics were not represented in our experiment, while others like death investigation and toxicology were overrepresented. The dataset, therefore, needs more additions to fully mirror the range or randomness of a real forensic case. Future research may also incorporate additional modalities in testing datasets such as audio and video to better benchmark the role of MLLMs in real-world forensics cases, which often involve dealing with evidence like gunshot recordings and CCTV camera footage ^38–40^.

Even with these constraints, it should be noted that this work represents novelty and value. We acknowledge the need to create better evaluations that reflect how foundation models would perform in the real world by including not just multiple choice questions, as was done with past medical benchmarks, but also including extended multi-party case study questions that require thorough reasoning in addition to immediate factual recall ^31–33,41^. By providing a benchmark for forensic science that incorporates modes of delivery motivated by an advancement toward realistic usage, we provide a foundation which will spur further meaningful development within this specialized field. The potential advantages of using AI in forensic practice are immense, adding to medical and scientific know-how and augmenting legal process and social impact ^10– 12,15,42^. This study lays critical groundwork for additional research to redesign forensic operations, increase decision-making, and help justice and public security.

Our evaluation revealed interpretable failure modes that point to clear directions of how MLLMs can be improved for them to meaningfully contribute to forensic tasks. Most models struggled with visual reasoning, specifically nuanced interpretation of forensic photographs. Others stumbled over open-ended queries, producing boilerplate responses or misinterpreting surface-level details. These issues hint at vulnerabilities in visual encoding, context retention, and multi-step inference—skills critical for application in forensics. Identification of these vulnerabilities directs future efforts toward model grounding, cross-modal correspondence, and case-specific inference improvement ^43–46^.

Better performance on multiple-choice items suggests models may be overfitting such in-training patterns common to testing conditions, such as those found on standard tests ^31–33^.

Generalizability to unstructured or novel forensic scenarios is thus made more difficult. Future benchmarking evaluations will need to include more open-ended tasks with imperfect information, better emulating actual challenges faced in forensic medicine.

Existing MLLMs can be potentially useful instructional tools for forensic science. Their strengths in systematic multiple-choice and text-based tasks can assist in concept reinforcement, exam preparation, and guided review ^5,47,48^. Their weaknesses, however, exclude independent and autonomous use in actual forensic medicine workspaces at the current moment. Their uses in live procedures need to be strictly in combination with human experts. They may assist in hypothesis testing, documentation, or case comparison but they cannot be fully authoritative help tools. Current limitations of foundation models in exercising full judgment, nuance, and ethical accountability preclude meaningfully realistic forensic decision-making ^49^.

Overall, state-of-the-art MLLMs are constrained in forensic reasoning but hold clear promise in improving their performance. Overcoming challenges in visual understanding and open-ended reasoning will require the development of more extensive, multimodal forensic datasets and domain-targeted pre- and post-training methods like fine-tuning ^44^. Uncertainty quantification, structured explanation, and task-aware prompting should also remain near-term priorities in advancing the reliability and transparency of model output ^50,51^. Through continued collaboration between forensic experts and machine learning experts, the models can potentially be of help in education, preliminary case screening, and administrative tasks. While not yet ready for standalone use, their combination with regulated human oversight could eventually increase the accuracy, uniformity, and availability of forensic operations.

## Data Availability

All data produced will be available upon publication, or reasonable request to the authors

## Notes

### Competing Interest Statement

The authors have declared no competing interest.

### Funding Statement

This study did not receive any funding

